# Ultrasound evaluation of vagus nerve cross-sectional area in a community-dwelling elderly Japanese cohort

**DOI:** 10.1101/2023.01.06.23284262

**Authors:** Kazumasa Oura, Hiroshi Akasaka, Naoki Ishizuka, Yuriko Sato, Masahiro Kudo, Takashi Yamaguchi, Mao Yamaguchi Oura, Ryo Itabashi, Tetsuya Maeda

## Abstract

**Objectives:** Although the vagus nerve (VN) is easily observed by ultrasonography, few studies have evaluated the cross-sectional area (CSA) of the VN in healthy older individuals from East Asia. In this study, we aimed to report reference values for the CSA of the VN in community-dwelling elderly Japanese individuals and to identify any associated medical history and/or lifestyle factors.

**Methods:** The present study included 336 participants aged ≥ 65 years from a prospective cohort study conducted in Yahaba, Japan from October 2021 to February 2022. The CSA of the VN was measured bilaterally at the level of the thyroid gland by ultrasonography. Univariate and multivariable linear regression analyses were conducted to identify the associations between clinical and background factors and the CSA of the VN on each side.

**Results:** In our cohort, the median CSA of the VN was 1.3 mm^2^ (interquartile range [IQR] 1.1– 1.6) on the right side and 1.2 mm^2^ (IQR 1.0–1.4) on the left side. Multivariable linear regression analysis showed that history of head injury (β = -0.15, *p* < .01), history of convulsion (β = 0.19, *p* < .01), and BMI (β = 0.30, *p* < .01) were independently associated with the CSA of the VN on the left side. In contrast, there were no independent associations between any of the assessed variables and the CSA on the right side.

**Conclusion:** We have reported reference VN CSA values for community-dwelling elderly Japanese individuals. In addition, we showed that the CSA of the VN on the left side was positively associated with a history of convulsive seizure and BMI and inversely associated with a history of head injury.

## Introduction

The utility of ultrasonography for imaging the vagus nerve (VN) was first reported in 1998 [1]. Since then, ultrasonography has been used to assess the cross-sectional area (CSA) of the VN in patients with a variety of neurological conditions in order to investigate potential pathological changes. Such conditions include neurodegenerative diseases such as Parkinson’s Disease [2–7] and amyotrophic lateral sclerosis [8–10], neuropathies such as Charcot-Marie-Tooth [11, 12], Guillain-Barré syndrome [13], and transthyretin familial amyloid polyneuropathy [14], as well as fibromyalgia [15]. Significant reductions in the CSA of the VN have been observed on the right side [5] or on both sides in Parkinson’s disease [3, 4, 6] and on both sides in ALS [9, 10], correlating variably with disease severity and duration and age [3, 4, 9, 10]. However, not all studies have identified differences [2, 7, 8]. In addition, Liu et al. [13] showed enlargement of the VN on both sides in a cohort of northern Chinese patients with Guillain-Barré syndrome subtypes; the VN was larger in patients exhibiting autonomic dysfunction. Furthermore, recently, we have shown the cross-sectional area (CSA) of the VN on the right side to be an independent predictor of atrial fibrillation (AF) in a cohort of Japanese patients following acute ischemic stroke or transient ischemic attack [16].

As ultrasound of the peripheral nerves is being increasingly used to conduct structural assessments in various diseases, well-defined references values for the CSA of the VN are essential. Recently, a meta-analysis of CSA reference values for the VN in healthy adults was published [17]. In addition, some studies have shown the VN CSA to be relatively small in Chinese patient cohorts; however, these studies were small-scale [14] or did not distinguish between left and right VN [13, 14] and were not conducted in particularly aged populations. Thus, overall, data regarding the VN in older East Asian—and specifically Japanese— populations are lacking. This study aimed to determine reference values for the CSA of the VN in community-dwelling elderly Japanese individuals and to identify any medical history and/or lifestyle factors associated with these values.

## Materials and Methods

### Study population

The Japan Prospective Studies Collaboration for Aging and Dementia (JPSC-AD) is a multicenter prospective cohort study of dementia that surveyed more than 10,000 community-dwelling elderly persons aged 65 years or older at eight study sites in Japan [18]. Among the participants in the JPSC-AD, only those who enrolled in the Yahaba Active Aging and Healthy Brain (YAHABA) study for the residents of Yahaba Town (Iwate Prefecture, Japan) were included in the present study. The YAHABA study is a community-based prospective cohort study that was established in 2016 to clarify the risk factors and etiology of dementia, cerebrovascular diseases, and movement disorders in older adults [19]. The study protocol complied with the ethical guidelines of the 2013 Declaration of Helsinki. The Iwate Medical University School of Medicine Institutional Ethics Committee reviewed and approved the protocol (no. HG2020-017), and written informed consent was obtained from all participants.

This cross-sectional study of the VN was conducted from October 2021 to February 2022. All participants underwent ultrasonography to evaluate the CSA of the VN. At the time of examination, the following data were collected: history of stroke, cardiovascular disease, congestive heart failure, AF, malignancy, respiratory disease, liver disease, convulsion, depression, head or non-head traumatic injury (i.e., fractures or injuries requiring hospitalization), Parkinson’s disease, and other neurological diseases (including dementia); the incidence of autonomic disorders in the subjects was not recorded. Moreover, clinical backgrounds including the prevalence of hypertension, dyslipidemia, diabetes mellitus (DM), and smoking and drinking habits were collected from all participants. Cardiovascular disease was defined as coronary heart disease or coronary intervention. Participants’ medical records and documents used for regular health checks in the YAHABA study were obtained and their medical history and biomarkers were compiled. All information on medical history was based on the self-reports of the study participants. Convulsion was defined as the previous occurrence of at least one convulsive seizure, with or without a diagnosis of epilepsy. Height, weight, body mass index (BMI), and blood pressure were measured. Blood pressure was measured three times using an automated sphygmomanometer with the study participants in a seated position after at least 5 minutes of rest; the average of the three measurements was calculated [18]. Height and weight were measured while the participants were wearing light clothing without shoes, and BMI was calculated. Casual blood samples were drawn from the antecubital vein of all participants to test the fasting blood glucose and hemoglobin A1c concentrations.

Participants with a fasting blood glucose concentration ≥ 126 mg/dL (7.0 mmol/L) or hemoglobin A1c concentration ≥ 6.5% or who reported a history of DM were considered to have DM. Although the YAHABA study examined other parameters such as liver and kidney function and lipid concentrations [19], only fasting blood glucose and hemoglobin A1c concentrations were analyzed in the present study.

### Ultrasound imaging

Ultrasound to evaluate the CSA of the VN was performed using an EPIQ CVx (Philips Japan, Tokyo, Japan), Aplio 400 (Canon Medical Systems, Otawara, Japan), Logiq S8 (GE Healthcare Japan, Tokyo, Japan), or Voluson E8 (GE Healthcare Japan, Tokyo, Japan). Linear probes (7– 12 MHz) were used in all examinations. Ultrasonography and measurements of the CSA of the VN were performed by one of the five medical laboratory technicians who specialized in ultrasound examination and were blinded to each patient’s clinical information; imaging was conducted once per patient. Cross-sectional imaging of the VN was recorded bilaterally at the level of the thyroid gland [2]. Measurements taken at this level have the advantages of easy identification and good inter-rater agreement [20], while analysis at a single position was used to minimize testing time and patient burden. Based on the Digital Imaging and Communications in Medicine files stored in the ultrasound equipment, the CSA (reported in mm^2^ by the ultrasound system) of the VN was measured after manual tracing of the periphery of the VN section.

### Statistical analysis

The values were expressed as median and interquartile range (IQR), mean and standard deviation, or number and frequency, as appropriate. Simple linear regression analysis was used to evaluate the associations between the CSA of the VN and the individual background characteristics of the participants. Factors that were significant in the simple linear regression analysis were selected for evaluation as independent parameters by multivariate analysis. For the multivariable linear regression analysis, the World Health Organization BMI classification was used to categorize the participants as underweight (BMI < 18.5 kg/m^2^), normal (BMI 18.5– 24.9 kg/m^2^), overweight (BMI 25.0–29.9 kg/m^2^), or obese (BMI ≥ 30 kg/m^2^) [21]. All statistical analyses were conducted using SPSS version 26 (IBM Japan, Tokyo, Japan), and *p* < 0.05 was considered statistically significant.

### Results

After excluding 10 subjects who were incapable of providing informed consent, a total of 336 participants were included in this study. The characteristics of the participants are shown in Table 1. Ultrasound imaging (Fig.1) showed that in our study population, the median CSA of the VN was significantly smaller on the left side than on the right side (1.2 mm^2^ versus 1.3 mm^2^, *p* < .01), in line with previous studies [17, 20, 22, 23].

**Table 1.** Characteristics of the study participants.

| Variables |  |
| --- | --- |
| Age (years), median [IQR] | 75.5 [73–80] |
| Male sex, no. (%) | 150 (44.6) |
| Stroke, no. (%) | 24 (7.1) |
| Cardiovascular disease, no. (%) | 20 (6.0) |
| Atrial fibrillation, no. (%) | 24 (7.1) |
| Congestive heart failure, no. (%) | 7 (2.1) |
| Malignancy, no. (%) | 51 (15.2) |
| Respiratory disease, no. (%) | 51 (15.2) |
| Liver disease, no. (%) | 43 (12.8) |
| Head injury, no. (%) | 27 (8.0) |
| Non-head injury, no. (%) | 124 (36.9) |
| Depression, no. (%) | 4 (1.2) |
| Convulsion, no. (%) | 3 (0.9) |
| Parkinson's disease, no. (%) | 0 (0) |
| Other neurological disease, no. (%) | 30 (8.9) |
| Hypertension, no. (%) | 218 (64.9) |
| Dyslipidemia, no. (%) | 136 (40.5) |
| Diabetes mellitus, no. (%) | 59 (17.6) |
| Current smoker, no. (%) | 108 (32.1) |
| Regular alcohol consumption, no. (%) | 128 (38.1) |
| Height (cm), median [IQR] | 154 [148–162] |
| Weight (kg), median [IQR] | 57 [49–66] |
| BMI (kg/m<sup>2</sup>), median [IQR] | 23.8 [21.6–26.2] |
| BMI category, no. (%) |  |
| <18.5 kg/m<sup>2</sup> | 14 (4.2) |
| 18.5–24.9 kg/m<sup>2</sup> | 199 (59.6) |
| 25.0–29.9 kg/m<sup>2</sup> | 102 (30.5) |
| ≥ 30 kg/m<sup>2</sup> | 19 (5.7) |
| Fasting blood glucose (mmol/L), median [IQR] | 6.53 [5.60–7.90] |
| Hemoglobin A1c (NGSP %), median [IQR] | 5.6 [5.4–5.9] |
| Systolic blood pressure (mmHg), median [IQR] | 143.7 [131.1–159.3] |
| Diastolic blood pressure (mmHg), median [IQR] | 75.0 [68.0–82.3] |
| Right VN CSA (mm<sup>2</sup>), median [IQR] | 1.3 [1.1–1.6] |
| Left VN CSA (mm<sup>2</sup>), median [IQR] | 1.2 [1.0–1.4] |

VN, vagus nerve; CSA, cross-sectional area; IQR, interquartile range; no., number; BMI, body
mass index; NGSP, National Glycohemoglobin Standardization Program.

**Fig 1.**
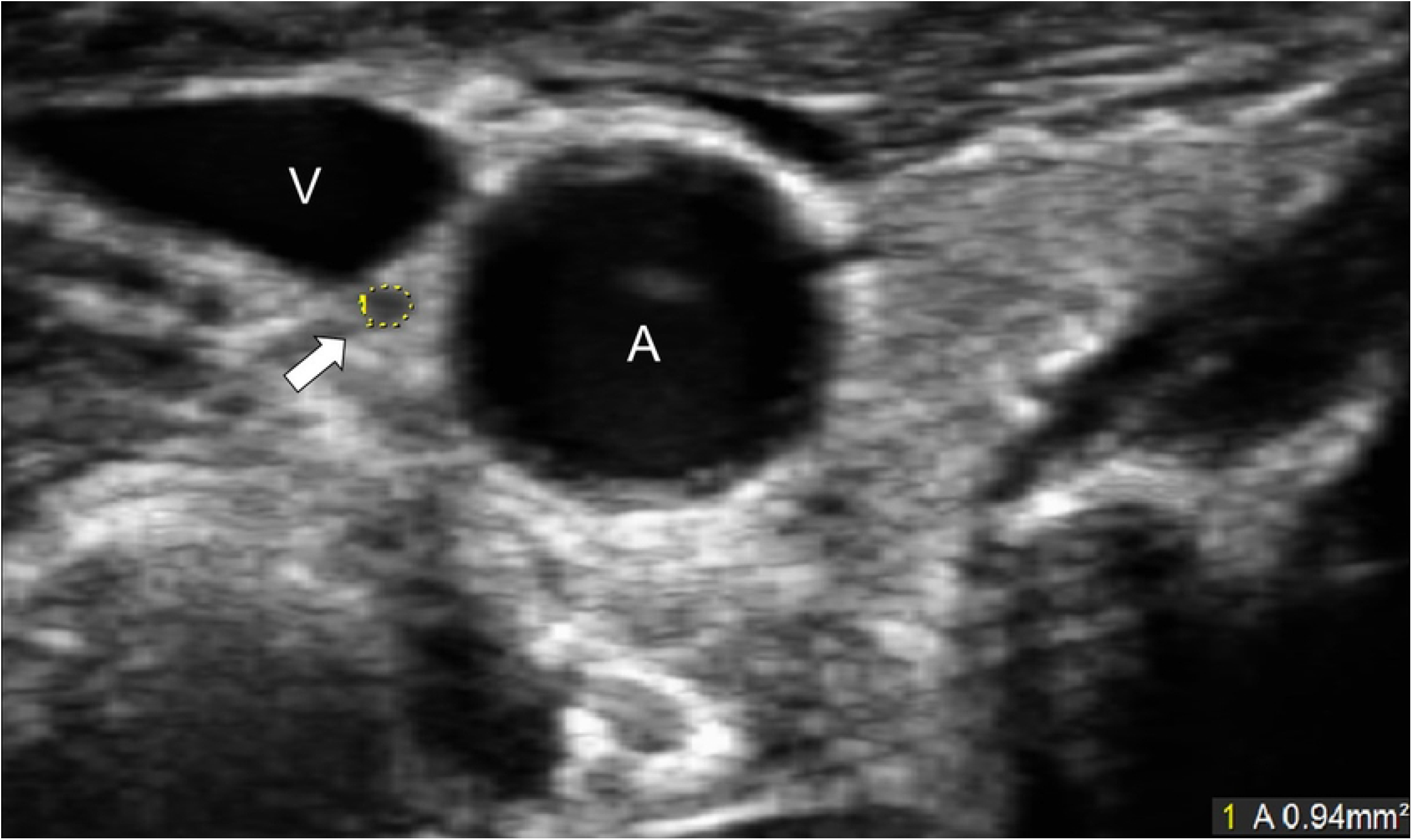
Ultrasonographic image of the vagus nerve. The vagus nerve is shown as a small, rounded, hypoechoic structure between the carotid artery and jugular vein. The cross-sectional area of the vagus nerve was measured by manual tracing (yellow circle). A, common carotid artery; V, jugular vein.

We conducted simple linear regression analysis to determine the relationship between the background of the participants and the CSA of the VN on the left and right sides (Table 2). Sex, height, body weight, BMI, and smoking habit were significantly associated with the CSA of the right VN. Furthermore, body weight, BMI, head injury, convulsion, hypertension, and smoking habit were significantly associated with the CSA of the left VN.

**Table 2.** Simple linear regression analysis of the association between vagus nerve cross-sectional area and background of the participants.

|  | Cross-sectional area of the vagus nerve |  |  |  |
| --- | --- | --- | --- | --- |
|  | Right |  | Left |  |
|  | R<sup>2</sup> | *p*-value | R<sup>2</sup> | *p*-value |
| Sex | 0.022 | < .01 | 0.006 | .15 |
| Age | 0.001 | .55 | 0.001 | .67 |
| Height | 0.036 | < .01 | 0.007 | .12 |
| Body weight | 0.040 | < .01 | 0.064 | < .01 |
| Body mass index | 0.015 | .03 | 0.067 | < .01 |
| Stroke | < 0.001 | .84 | < 0.001 | .95 |
| Cardiovascular disease | 0.004 | .26 | < 0.001 | .73 |
| Atrial fibrillation | 0.001 | .50 | < 0.001 | .94 |
| Congestive heart failure | < 0.001 | .78 | < 0.001 | .99 |
| Malignancy | < 0.001 | .84 | < 0.001 | .69 |
| Respiratory disease | < 0.001 | .87 | 0.001 | .54 |
| Liver disease | 0.003 | .35 | 0.001 | .54 |
| Head injury | 0.005 | .21 | 0.014 | .03 |
| Non-head injury | < 0.001 | .82 | 0.002 | .43 |
| Depression | < 0.001 | .92 | 0.002 | .37 |
| Convulsion | 0.002 | .42 | 0.025 | < .01 |
| Other neurological disease | 0.002 | .40 | < 0.001 | .97 |
| Hypertension | 0.002 | .41 | 0.012 | .04 |
| Dyslipidemia | 0.002 | .43 | < 0.001 | .94 |
| Diabetes mellitus | < 0.001 | .98 | 0.001 | .53 |
| Fasting blood glucose | 0.005 | .21 | < 0.001 | .72 |
| Hemoglobin A1c | 0.001 | .60 | 0.001 | .61 |
| Current smoker | 0.013 | .03 | 0.013 | .04 |
| Regular alcohol consumption | 0.008 | .11 | .003 | .30 |
| Systolic blood pressure | < 0.001 | .74 | 0.005 | .18 |
| Diastolic blood pressure | 0.001 | .52 | 0.005 | .21 |

Multivariable linear regression analysis with adjustments for age, sex, BMI, AF, DM, head injury, convulsion, hypertension, and smoking habit showed that history of head injury (β = - 0.15, *p* < .01), history of convulsion (β = .19, *p* < .01), and BMI (β = .30, *p* < .01) were independently associated with the CSA of the VN on the left side, while none of the variables were significantly associated with the CSA of the VN on the right side (Table 3).

**Table 3.** Multivariable linear regression analysis of variables associated with vagus nerve cross-sectional area.

|  | Right cross-sectional area |  | Left cross-sectional area |  |
| --- | --- | --- | --- | --- |
| | $\beta$ | *p*-value | $\beta$ | *p*-value |
| Male sex | -0.13 | .10 | 0.05 | .50 |
| Age | 0.03 | .61 | 0.04 | .47 |
| Atrial fibrillation | -0.06 | .31 | 0.01 | .86 |
| Head injury | -0.08 | .15 | -0.15 | < .01 |
| Convulsion | 0.05 | .32 | 0.19 | < .01 |
| Hypertension | 0.02 | .79 | 0.02 | .74 |
| Current smoking habit | 0.02 | .80 | 0.11 | .13 |
| Diabetes mellitus | -0.01 | .83 | -0.004 | .94 |
| Body mass index | 0.06 | .28 | 0.30 | < .01 |

## Discussion

The present study provides reference values for the CSA of the VN in Japanese community-dwelling individuals aged 65 years and older. Moreover, multivariable linear regression analysis revealed that the CSA of the VN on the left side was positively associated with a history of convulsion and higher BMI and inversely associated with a history of head injury. In contrast, there were no independent associations between any of the assessed variables and the CSA of the VN on the right side.

The present study measured the CSA of the VN by ultrasound in a relatively large cohort, compared with previous studies. CSA values for the VN measured by ultrasound in healthy individuals vary widely among different studies, ranging from 1.3 to 6.0 mm^2^ on the right side and from 1.1 to 5.9 mm^2^ on the left side [2–5, 7–12, 15, 20, 22–25] (see Table 4). Since the methodology (ultrasound and resolution) for all these studies were the same, this discrepancy between studies may be explained by differences in the study cohorts. Most previous studies have been conducted in non-Asian countries; however, the CSA values obtained in our study appeared comparable to those found in a small number of Chinese studies [12–14]. For example, the median CSA of the left VN in a pilot study conducted by Du et al. [14] was 1.0 mm^2^ (range 1.0–2.0). Moreover, the participants in our study were older than previously studied cohorts, as only participants aged 65 years or older were included. Pelz et al. reported that the CSA of the VN was inversely correlated with age [20], although another study, conducted by Laucius et al., failed to find any differences in VN CSA between younger and older age cohorts [26]; however, the maximum age of the subjects in the latter study was 55 years. Thus, the relatively small CSA of the VN observed in the present study may have been related to the advanced age of the participants. In addition, the somewhat unexpected lack of association between VN CSA and age in our study may have been because in individuals who are already relatively old (≥ 65 years), the CSA of the VN may be unaffected by further aging.

**Table 4.** Results of current and previous ultrasound studies describing the cross-sectional area of the vagus nerve in healthy individuals.

| First author's name | Number of subjects | Age (years), mean (SD) | Height (cm), mean (SD) | Weight (kg), mean (SD) | BMI (kg/m<sup>2</sup>), mean (SD) | Right CSA (mm<sup>2</sup>), mean (SD) | Left CSA (mm<sup>2</sup>), mean (SD) |
| --- | --- | --- | --- | --- | --- | --- | --- |
| Present study | 336 | 76.9 (5.5) | 155.0 (9.2) | 58.3 (11.9) | 24.1 (3.8) | 1.4 (0.4) | 1.2 (0.4) |
| Sijben LCJ [7]<sup>a</sup> | 51 | 72 (8) | NA | NA | NA | 2.2 (0.7) | 1.9 (0.6) |
| Papadopoulou M [9]<sup>a</sup> | 28 | 57 (8.9) | NA | NA | 27.3 (4.6) | 2.9 (0.8) | 2.3 (0.8) |
| Papadopoulou M [15]<sup>a</sup> | 28 | 50 (NA) | NA | NA | 25.4 (NA) | 3.2 (0.8) | 2.7 (1.1) |
| Liu L [13]<sup>b</sup> | 40 | 50.8 (12.8) | 167.6 (7.8) | 65.4 (10.6) | NA | 1.3 (0.5) | NA |
| Horsager J [5]<sup>a</sup> | 56 | 69.1 (8.9) | NA | NA | 24.9 (3.7) | 2.4 (0.6) | 2.0 (0.5) |
| Sartucci F [25]<sup>a</sup> | 20 | 65.2 (10.3) | 169.9 (7.1) | 71.0 (16.9) | 24.5 (5.0) | 6.0 (1.3) | 5.6 (1.3) |
| Holzapfel K [10]<sup>a</sup> | 19 | 63.1 (11.1) | NA | NA | NA | 2.2 (0.6) | 2.0 (0.3) |
| Curcean AD [22]<sup>a</sup> | 21 | 25 (2) | 174.0 (10.0) | 72.0 (17.0) | 23.6 (4.4) | 2.9 (1.3) | 2.1 (0.8) |
| Walter U [23]<sup>a</sup> | 70 | M 43.7 (20.3)<br>F 45.5 (19.9) | 173.6 (7.9) | 74.6 (15.7) | NA | M 1.8 (0.6)<br>F 2.0 (0.7) | M 1.4 (0.5)<br>F 1.6 (0.7) |
| Pelz JO [3]<sup>a</sup> | 35 | 67.9 (6.2) | 167.7 (9.1) | 74.9 (14.4) | NA | 2.5 (0.5) | 1.8 (0.4) |
| Pelz JO [20]<sup>a</sup> | 60 | 49.7<br>(19.7) | 170.0<br>(9.0) | 71.0<br>(13.0) | NA | 2.6 (0.6) | 1.9 (0.4) |
| Walter U [4]<sup>a</sup> | 61 | 45.1<br>(20.7) | 173.8<br>(8.1) | 75.1<br>(16.8) | NA | 1.3 (0.5) | 1.1 (0.5) |
| Fedtke N [2]<sup>a</sup> | 15 | NA | NA | NA | NA | 2.7 (0.7) | 2.4 (0.7) |
| Cartwright MS<br>[11]<sup>c</sup> | 24 | 38.6<br>(NA) | NA | NA | NA | NA | 5.9 (NA) |
| Cartwright MS<br>[24]<sup>c</sup> | 60 | 45.9<br>(NA) | 168.0<br>(NA) | 74.5<br>(NA) | 26.5<br>(NA) | 5.0 (2.0) | NA |
| Weise D [8]<sup>a</sup> | 40 | 65.7<br>(11.8) | 171.0<br>(9.0) | 75.3<br>(12.1) | 25.8<br>(3.8) | 1.6 (0.5) | 1.4 (0.5) |
| Du K [14]<sup>b</sup> | 17 | 47.6<br>(12.8) | 170<br>(7.0) | 66.5<br>(6.5) | 22.0<br>(1.1) | 2.0<sup>†</sup><br>(1.0–<br>3.0)<sup>#</sup> | 1.0<sup>†</sup><br>(1.0–2.0)<sup>#</sup> |
| Niu J [12]<sup>b</sup> | 105 | 42<br>(16.1) | 167.2<br>(8.3) | 65<br>(10.8) | 23.2<br>(2.8) | 1.0<sup>†‡</sup><br>(1.0–<br>2.0)<sup>#</sup> |  |

VN, vagus nerve; CSA, cross-sectional area; SD, standard deviation; NA, not available; M,
Male; F, Female. Studies were conducted in Europe<sup>a</sup>, China<sup>b</sup>, and the US<sup>c</sup>. <sup>†</sup> median value; <sup>#</sup>
range; <sup>‡</sup> a single value for the vagus nerve was given.

Our findings indicated that the CSA of the VN on the left side was associated with BMI. Although an association between BMI and VN CSA is to be expected, some previous reports have indicated a lack of correlation between weight and VN CSA on either side [20, 22, 23]. The lower BMI of our study cohort, compared with previous study cohorts, may also have contributed to the smaller VN CSA in our study. Furthermore, our study found no association between the CSA of the VN and DM, despite a previous study showing that the functional status of the VN was reduced in DM patients [27].

We showed that the CSA of the VN on the left side was significantly smaller in participants with a history of head injury than in those without such a history. Interestingly, history of non-head injury was not associated with the CSA of the VN. The smaller CSA of the left VN in those with prior head injury may be related to autonomic failure. However, this is necessarily somewhat speculative as the exact nature of the head injuries and resulting brain damage was not ascertained in the study participants; further studies are required to verify this association. Traumatic head injury can be caused by orthostatic hypotension, which is a typical symptom of autonomic failure [28]. Head injury due to orthostatic hypotension is especially frequent in the elderly population [29], and orthostatic hypotension is significantly positively associated with falls in older adults [30]. Heart rate variability can be used to assess autonomic nervous system function and is reportedly inversely associated with the CSA of the left VN [31]. Furthermore, atrophy of the VN has been reported in patients with Parkinson’s disease [32] and has been observed to correlate with parasympathetic dysfunction [6]. However, other studies have contradicted these findings [2, 7]. Although none of the present study participants had neurological disorders featuring autonomic neuropathy, we are currently conducting a study in patients with Parkinson’s disease to investigate the usefulness of CSA of the VN with regard to diagnosis.

In our cohort, a small percentage of participants had suffered at least one convulsive seizure. As these were older individuals, some of these patients may have been epileptic without having been formally diagnosed. Interestingly, although the relationship between convulsions and CSA of the VN is currently unknown, we showed that the CSA of the VN on the left side was significantly larger in individuals with a history of convulsive seizure. VN stimulation is known to be an effective treatment for epilepsy, reducing the frequency and intensity of seizures [33]. The suggested mechanisms include afferent vagal projections to seizure-generating regions in the basal forebrain and insular cortex [34], effects on the locus ceruleus [35,36], desynchronization of hypersynchronized cortical activity [37, 38], and cortical inhibition secondary to the release of inhibitory neurotransmitters [39, 40]. Nonetheless, the precise mechanisms by which VN stimulation therapy achieves seizure reduction have not been fully elucidated. Assuming that the CSA of the VN on the left side might be spontaneously enlarged to control seizures, greater CSA might be a consequence rather than a cause of epilepsy. As this possibility requires further investigation, in future studies we aim to measure the CSA of the VN in patients admitted with convulsive seizures.

The present study had some limitations. First, the accuracy of the CSA measurements was not examined. Five medical laboratory technicians measured the CSA of the VN in our study cohort, but intra- and inter-observer reliabilities could not be determined as each CSA was measured only once by a single technician. Second, as this was a cross-sectional study, we could not address causal relationships between the variables and the CSA of the VN. Third, the resolution of the ultrasound probes was relatively low for the measurement of nerves; future studies should use probes with resolutions of 15 MHz or more. Fourth, as medical history (including previous injuries) was self-reported by the study participants, there was potential for inaccuracies in the information recorded. Finally, in the future, it will be important to evaluate the CSAs of nerves other than the VN in a Japanese cohort, to see if they are also smaller than the values reported for other populations.

In conclusion, we determined reference values for the CSA of the VN in a large cohort of community-dwelling elderly Japanese individuals. The CSA of the VN on the left side was positively associated with history of convulsive seizure and BMI and inversely associated with history of head injury. In contrast, there were no independent associations between any of the assessed variables and the CSA on the right side. A longitudinal prospective cohort study is warranted to evaluate the causal relationships between the clinical/background factors identified in this study and the CSA of the VN.

## Data Availability

All relevant data are within the manuscript and its Supporting Information files.

## Acknowledgments

We thank Kelly Zammit, BVSc, from Edanz (https://jp.edanz.com/ac) for editing a draft of this manuscript and Sarah Ivins from Edanz (https://jp.edanz.com/ac) for editing and medical writing support.

## Notes

### Competing Interest Statement

The authors have declared no competing interest.

### Funding Statement

This study was supported by Grants-in-Aid from the Japan Agency for Medical Research and Development (JP21dk02070532). The funder had no role in study design, data collection and analysis, decision to publish, or preparation of the manuscript.

### Author Declarations

The Iwate Medical University School of Medicine Institutional Ethics Committee reviewed and approved the protocol (no. HG2020-017), and written informed consent was obtained from all participants.

